# Profiling placental DNA methylation associated with maternal SSRI treatment during pregnancy

**DOI:** 10.1101/2022.06.21.22276723

**Authors:** Amy M. Inkster, Chaini Konwar, Maria S. Peñaherrera, Ursula Brain, Almas Khan, E. Magda Price, Johanna M. Schuetz, Élodie Portales-Casamar, Amber Burt, Carmen J. Marsit, Cathy Vaillancourt, Tim F. Oberlander, Wendy P. Robinson

## Abstract

**Background:** Selective serotonin reuptake inhibitors (SSRIs) for treatment of prenatal maternal depression have been associated with neonatal neurobehavioral disturbances, though the molecular mechanisms remain poorly understood. *In utero* exposure to SSRIs may affect DNA methylation (DNAme) in the human placenta, an epigenetic mark that is established during development and is associated with gene expression.

**Methods:** Chorionic villus samples from 64 human placentas were profiled with the Illumina MethylationEPIC BeadChip; clinical assessments of maternal mood and SSRI treatment records were collected at multiple time points during pregnancy. Case distribution was 20 SSRI-exposed cases and 44 SSRI non-exposed cases. Maternal depression was defined using a mean maternal Hamilton Depression score >8 to indicate symptomatic depressed mood (“maternally-depressed”), and we further classified cases into SSRI-exposed, maternally-depressed (n=14); SSRI-exposed, not maternally-depressed (n=6); SSRI non-exposed, maternally-depressed (n=20); and SSRI non-exposed, not maternally-depressed (n=24). For replication, Illumina 450K DNAme profiles were obtained from 34 additional cases from an independent cohort (n=17 SSRI-exposed, n=17 SSRI non-exposed).

**Results:** No CpGs were differentially methylated at FDR < 0.05 comparing SSRI-exposed to non-exposed placentas, in a model adjusted for mean maternal Hamilton Depression score, or in a model restricted to maternally-depressed cases with and without SSRI exposure. However, at a relaxed threshold of FDR < 0.25, five CpGs were differentially methylated (|Δβ| > 0.03) by SSRI exposure status. Four were covered by the replication cohort measured by the 450K array, but none replicated. No CpGs were differentially methylated (FDR < 0.25) comparing maternally depressed to not depressed cases. In sex-stratified analyses for SSRI-exposed versus non-exposed cases (females n=31; males n=33), three additional CpGs in females, but none in males, were differentially methylated at the relaxed FDR < 0.25 cut-off.

**Conclusions:** We did not observe large-scale alterations of DNAme in placentas exposed to maternal SSRI treatment compared to placentas with no SSRI exposure. We also found no evidence for altered DNAme in maternal depression-exposed versus depression non-exposed placentas. This novel work in a prospectively-recruited cohort with clinician-ascertained SSRI exposure and mood assessments would benefit from future replication.

## BACKGROUND

As many as 20% of pregnant individuals suffer from mood disorders such as depression during pregnancy, and up to 6% of all pregnant individuals are treated with antidepressants such as selective serotonin reuptake inhibitors (SSRIs), based on estimates from several North American and European studies (1–4). Clinical management decisions for treatment of perinatal depression are complex, and in each case the clinician must balance the risks of treatment with the severity of depressive symptoms (5). Given the prevalence of maternal depression and SSRI treatment during gestation, much research has been conducted focusing on the fetal effects of both (6,7).

Maternal prenatal depressed mood has been associated with preterm birth (< 37 weeks of gestation), growth and developmental delays, and increased postnatal infant stress (8,9). Additionally, the literature suggests that the presence of maternal depressive symptoms independent of SSRI therapy is associated with increases in offspring childhood internalizing behaviours and affective disorders, though these exposures are hard to separate in human studies (10). Animal studies have permitted further investigation into the effect of maternal depressed mood on offspring outcomes by enabling the randomization of SSRI treatment. Rodent models have confirmed that associations identified between maternal depressed mood and offspring anxiety disorders are not mediated by SSRI exposure, and that SSRI treatment does not protect offspring from these outcomes; this topic is reviewed in (10).

Regarding the impacts of SSRI treatment on the feto-placental unit, Oberlander *et al*. previously found that maternal SSRI treatment was associated with altered blood flow to the fetus, increased rates of fetal hypoxemia, altered early neurodevelopment, lower arousal index throughout the newborn period, and altered psychomotor test scores (6,7,11–13). Prenatal SSRI exposure also tended to be associated with lower fetal-placental weight ratios and poorer fetal vascular perfusion, indicative of less efficient fetal blood flow to or from the placenta (14). Both of these factors indicate less efficient placental function in association with prenatal SSRI exposure, which may be related to downstream neonatal cognitive outcomes (14). Exposure to maternal SSRI treatment has previously been reported to affect syncytializaton of placental trophoblasts and extravillous trophoblast function, and to affect placental serotonin levels (15–17). Taken together, the effect of SSRI exposure on pregnancy outcomes suggests an interplay between the placenta and SSRI pharmacobiology, which could occur by a variety of mechanisms. It has, however, been postulated that both depression and SSRI exposure may impact fetal outcomes via altered serotonin signalling and downstream effects.

Serotonin (5-hydroxytryptamine, or 5-HT) is a central neurotransmitter and a critical signaling factor in many contexts, including during prenatal neurodevelopment of the fetus (18). Low levels of serotonin in the central nervous system have historically been associated with depressive symptoms in adults, in the 5-HT theory of depression (19). SSRI therapeutics, which increase extracellular levels of 5-HT, have been associated with alleviation of depressive symptoms (20). *In vivo*, serotonin is synthesized from the essential amino acid L-tryptophan, by tryptophan hydroxylase enzymes 1 and 2 (TPH-1, TPH-2), and is degraded by monoamine oxidases A and B (MAOA, MAOB) (21,22). SSRIs function to block the serotonin transporter (SERT, also known as 5-HTT (5-hydroxytryptamine transporter)), inhibiting the reabsorption of 5-HT by the presynaptic neurons and thereby increasing extracellular serotonin levels in the central nervous system (13). Several aspects of serotonin and SSRI biology are relevant to prenatal development, and both maternal depression and SSRI exposure have been associated with altered fetal outcomes. Notably, serotonin is first expressed early in gestation, when placental TPH-1 and TPH-2 convert maternal L-tryptophan to serotonin *de novo* (22,23). Over the course of gestation, serotonin (5-HT) is essential to many processes of prenatal development including embryogenesis, placentation, and neurodevelopment (21,24,25). SSRIs readily cross the placenta, leading to fetal drug exposure and altered fetal cardiac autonomic activity, presumably via altered serotonin signaling in the placenta and/or fetus (26,27).

One plausible mechanism by which SSRI exposure may impact pregnancy outcomes is via DNA methylation (DNAme). DNAme is an epigenetic mark involving the addition of a methyl group to the 5’ carbon of a cytosine molecule, which usually occurs in the context of cytosine-guanine dinucleotides (CpGs). In human tissues, DNAme signatures are associated with gene expression patterns (28), and are altered in association with certain environmental and chemical exposures, such as air pollution, arsenic, and tobacco smoke; for a review of this topic see (29).

DNAme patterns in the human placenta vary in association with certain maternal exposures and environments, but are relatively stable once established and serve as a possible record of gene expression patterns and exposures in gestation (30,31). The placenta exists at the interface of the maternal environment and the developing fetus, and environmental factors can gain access to the fetus in many cases only via the placenta (32,33). As SSRIs readily cross the placenta (34), it is possible that SSRI species alter placental DNAme patterns directly via association with epigenetic readers and writers (DNMT or TET enzymes), or through altering gene expression patterns in the serotonin signalling pathway, or other related pathways (29).

In this study, we hypothesized that prenatal SSRI treatment would be associated with distinct placental DNAme signatures. In a prospective cohort of 64 placentas with extensive maternal mood assessments during gestation, both with and without SSRI exposure, we investigated the effects of maternal SSRI treatment and maternal depressed mood on placental DNA methylation profiles. We considered the effect of SSRI exposure on placental DNAme as our primary outcome. The secondary outcome considered was the impact of maternal depression, which was investigated among participants not treated with SSRIs. Additionally, given previously reported sex differences in neurodevelopmental and placental outcomes related to maternal stress (35–40), we also assessed placental DNAme associated with maternal SSRI treatment in separate sex-stratified models.

## METHODS

### Discovery cohort

Participants were recruited as part of a prospective, longitudinal cohort approved by the University of British Columbia/Children’s and Women’s Health Centre of British Columbia research ethics board: H12-00733 (40). Written informed consent was obtained from all participants, and all procedures complied with the ethical standards on human experimentation and with the Helsinki Declaration of 1975 (revised in 2008). This specific study was additionally approved under certificate H16-02280.

Pregnant individuals with and without prenatal diagnoses of clinical depression, and with/without SSRI treatment plans were recruited at the British Columbia Women’s Hospital (BCWH) in the 20^th^ week of gestation (n=64). At recruitment, all pregnant individuals received the MINI assessment to screen for DSM-IV Axis I Depression. SSRI use was reported by the treating clinicians, and was defined as treatment for at least 75 days including the 3^rd^ trimester with one of the following: fluoxetine (Prozac), paroxetine (Paxil), sertraline (Zoloft), citalopram (Celexa), escitalopram (Lexapro), or venlafaxine (Effexor). Compliance with SSRI treatment was assessed and recorded at each prenatal study visit. While placental drug concentrations were not available at the time of this study, previous work on this cohort indicated that maternal plasma drug concentrations during gestation were consistent with self-report of SSRI use (26). Maternal mood was assessed at recruitment, 36 weeks of gestation, postnatal day 6, and at 24 months postnatally. The mood assessment included multiple clinician and patient-rated measures, of which the Hamilton Depression score, a 17-item clinician implemented assessment of the severity of depressive symptoms (41), and the Edinburgh Postnatal Depression score (EPDS), a 10-item self-assessment screening tool for depression (42), were analyzed.

Exclusion criteria were: bipolar illnesses, hypertension, any diabetes, current substance abuse, placental insufficiency, multi-fetal pregnancies, infants with congenital brain malformations, intrauterine growth restriction, or preterm birth. For all participants, measures of obstetric history, prenatal medication use, and sociodemographic variables were obtained. At birth, additional clinical information was collected for each infant including gestational age at delivery, infant sex assigned at birth, and infant birth weight; placentas were collected for DNAme studies.

After delivery, chorionic villus samples were obtained from 4 distinct cotyledonary sites (1.5-2 cm^3^) below the surface of the fetal-facing side of each placenta, as previously described (43). DNA was extracted from each of the 4 sites and pooled in equimolar amounts to provide a representative sample of the whole placenta prior to obtaining DNAme profiles. DNAme data for these 64 cases was collected in a single processing batch using the Illumina MethylationEPIC (“EPIC”) array platform (Illumina, San Diego, CA, USA), which profiles >850,000 CpGs genome-wide.

Case distribution was as follows: 20 SSRI-exposed and 44 SSRI non-exposed samples (Table 1). Cases were additionally sub-categorized into four groups for analysis based on both SSRI exposure and maternal depression status using a mean maternal Hamilton Depression score of >8 to indicate symptomatic depressed mood (“maternally depressed”) as follows: SSRI-exposed, maternally-depressed (n=14); SSRI-exposed without maternal depression (n=6); SSRI non-exposed, maternally depressed (n=20); and SSRI non-exposed without maternal depression (n=24), see Table 2 and Figure 1.

**Table 1.** Demographics of the discovery and replication cohorts by SSRI exposure status. SSRI refers to selective-serotonin reuptake inhibitor treatment during pregnancy. Birthweight Z-scores were calculated using gestational-age and sex-adjusted birthweight curves (45).

| **Discovery Cohort** |  |  |  |
| --- | --- | --- | --- |
|  | **SSRI-exposed** | **Non-SSRI-exposed** | ***p* value†** |
| **n** | 20 | 44 |  |
| **Maternal Hamilton Depression Score (mean (SD))** |  |  |  |
| 26 weeks | 10.6 (5.4) | 6.6 (3.6) | 0.001** |
| 36 weeks | 10.8 (4.2) | 8.3 (3.6) | 0.021* |
| Mean | 10.6 (4.5) | 7.5 (3.5) | 0.004** |
| **Length SSRI exposure (days, mean (SD))** | 264.4 (29.9) | - | - |
| **Type of SSRI exposure (n (%))** □ |  |  |  |
| Citalopram | 7 (35.0) | - | - |
| Escitalopram | 7 (35.0) | - |  |
| Fluoxetine | 1 (5.0) | - |  |
| Paroxetine | 1 (5.0) | - |  |
| Sertraline | 4 (20.0) | - |  |
| **Delivery type (n vaginal (%))** | 10 (50.0) | 29 (65.9) | 0.351 |
| **Gestational age at birth (weeks, mean (SD))** | 39.3 (1.4) | 39.6 (1.3) | 0.438 |
| **Gestational diabetes (n, mean (SD))** | 0 (0) | 0 (0) | - |
| **Infant sex (n male (%))** | 8 (40.0%) | 25 (56.8%) | 0.328 |
| **Infant birthweight Z-score (mean (SD))** | -0.01 (1.00) | -0.03 (0.73) | 0.915 |
| **PlaNET Ancestry<sup>§</sup> (mean (SD))** |  |  |  |
| Coordinate 1 | 0.00 (0.00) | 0.00 (0.01) | 0.354 |
| Coordinate 2 | 0.05 (0.21) | 0.13 (0.31) | 0.300 |
| Coordinate 3 | 0.95 (0.21) | 0.86 (0.31) | 0.286 |
| **Replication Cohort** |  |  |  |
|  | **SSRI-exposed** | **Non-SSRI-exposed** | ***p* value†** |
| **n** | 17 | 17 |  |
| **Type of SSRI exposure (n (%))** □ |  |  |  |
| Citalopram | 3 (17.6) | - | - |
| Escitalopram | 2 (11.8) | - |  |
| Fluoxetine | 2 (11.8) | - |  |
| Paroxetine | 1 (5.9) | - |  |
| Sertraline | 9 (52.9) | - |  |
| **Delivery type (n vaginal (%))** | 4 (23.5) | 12 (70.6) | 0.016* |
| **Gestational age at birth (weeks, mean (SD))** | 39.0 (0.9) | 39.0 (0.9) | 0.999 |
| **Infant sex (n male (%))** | 7 (41.2) | 10 (58.8) | 0.493 |
| **Gestational diabetes (n mean (SD))** | 5 (29.4) | 3 (18.8) | 0.758 |
| **PlaNET Ancestry<sup>§</sup>** |  |  |  |
| Coordinate 1 | 0.00 (0.01) | 0.08 (0.25) | 0.212 |
| Coordinate 2 | 0.01 (0.04) | 0.12 (0.32) | 0.174 |
| Coordinate 3 | 0.98 (0.05) | 0.80 (0.38) | 0.057 |

†p values are from chi-squared tests for categorical variables and t-tests for continuous variables,
comparing SSRI-exposed to SSRI non-exposed, \* indicates <0.05, \*\* indicates <0.005.
§The PlaNET algorithm outputs DNAME-based ancestry probability values ranging from 0-1,
which sum to 1 for each sample. Coordinate 1 is associated with probability of African ancestry,
coordinate 2 with East Asian ancestry, and coordinate 3 with European ancestry (46).

**Table 2.**
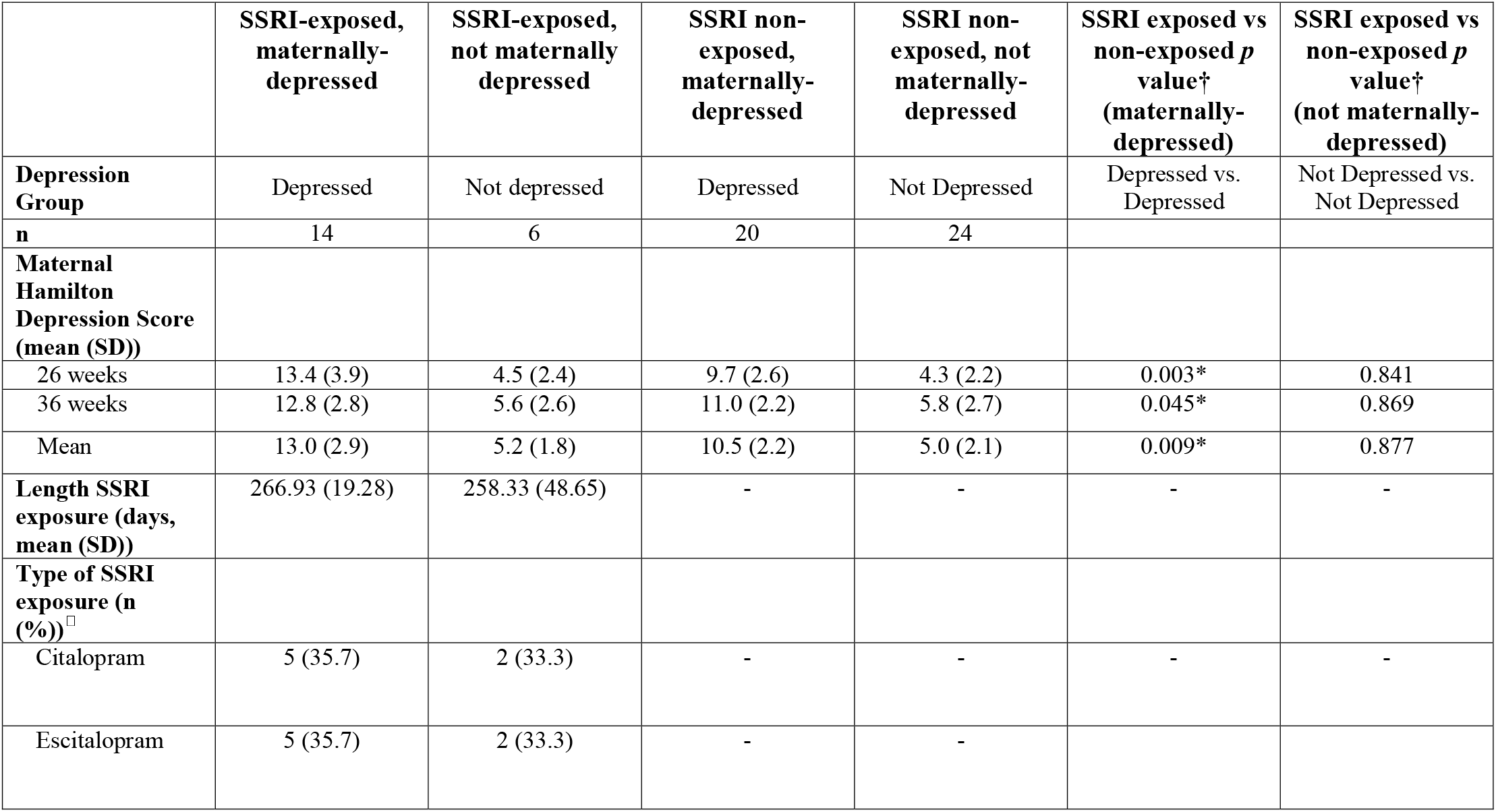
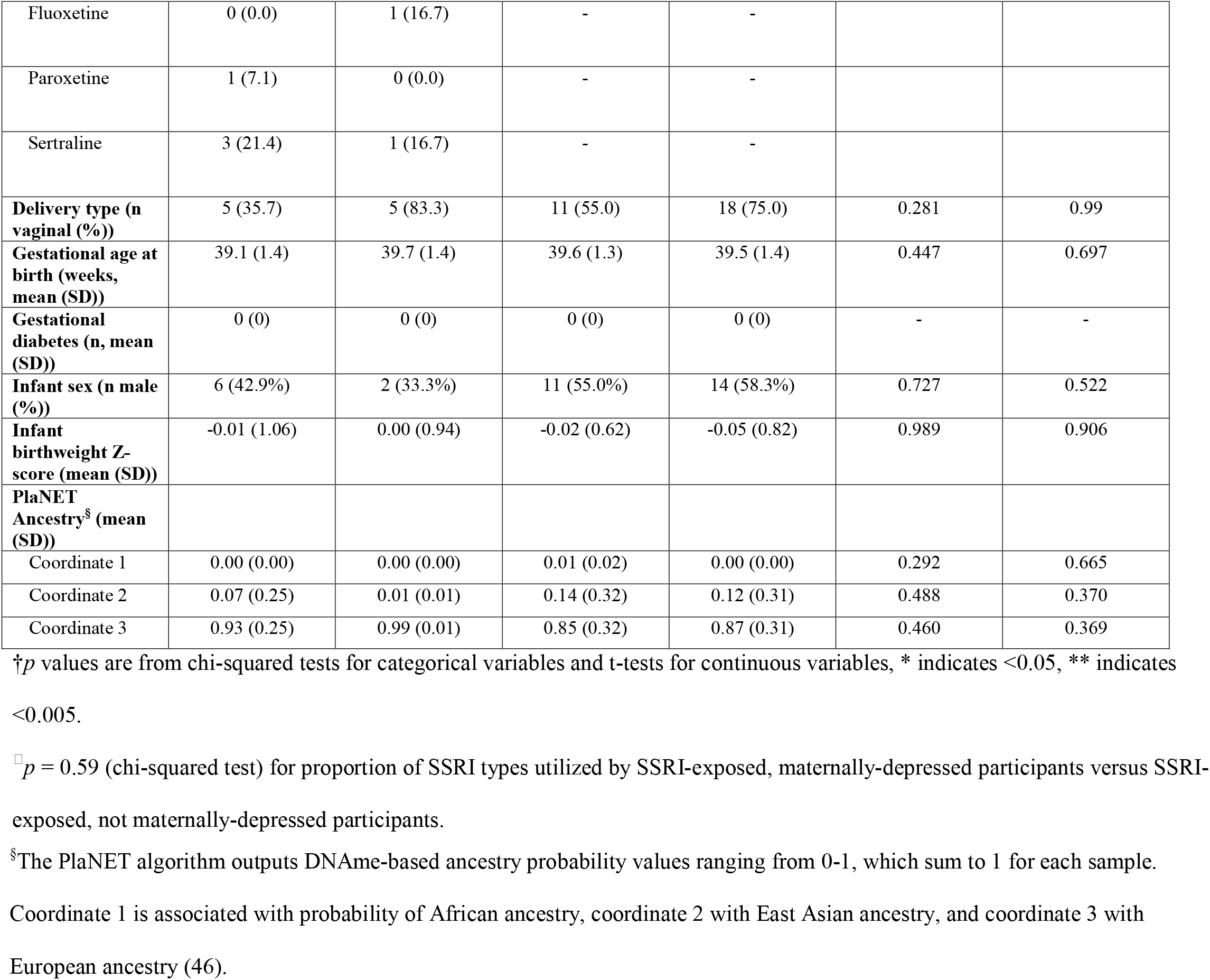
Discovery cohort demographics by SSRI and depression group status. Maternal depression was categorized using mean maternal Hamilton Depression score, < 8 was categorized as not depressed, > 8 as depressed, this is reflected in the “Depression Group” row. SSRI refers to selective-serotonin reuptake inhibitor treatment during pregnancy. Birthweight Z scores were calculated using gestational-age and sex-adjusted birthweight curves (45).

|  | SSRI-exposed,<br>maternally-depressed | SSRI-exposed,<br>not maternally<br>depressed | SSRI non-exposed,<br>maternally-depressed | SSRI non-exposed, not<br>maternally-depressed | SSRI exposed vs<br>non-exposed *p*<br>value<sup>†</sup><br>(maternally-depressed) | SSRI exposed vs<br>non-exposed *p*<br>value<sup>†</sup><br>(not maternally-depressed) |
| --- | --- | --- | --- | --- | --- | --- |
| **Depression**<br>**Group** | Depressed | Not depressed | Depressed | Not Depressed | Depressed vs.<br>Depressed | Not Depressed vs.<br>Not Depressed |
| **n** | 14 | 6 | 20 | 24 |  |  |
| **Maternal**<br>**Hamilton**<br>**Depression Score**<br>**(mean (SD))** |  |  |  |  |  |  |
| 26 weeks | 13.4 (3.9) | 4.5 (2.4) | 9.7 (2.6) | 4.3 (2.2) | 0.003* | 0.841 |
| 36 weeks | 12.8 (2.8) | 5.6 (2.6) | 11.0 (2.2) | 5.8 (2.7) | 0.045* | 0.869 |
| Mean | 13.0 (2.9) | 5.2 (1.8) | 10.5 (2.2) | 5.0 (2.1) | 0.009* | 0.877 |
| **Length SSRI**<br>**exposure (days,**<br>**mean (SD))** | 266.93 (19.28) | 258.33 (48.65) | - | - | - | - |
| **Type of SSRI**<br>**exposure (n**<br>**(%))<sup>‡</sup>** |  |  |  |  |  |  |
| Citalopram | 5 (35.7) | 2 (33.3) | - | - | - | - |
| Escitalopram | 5 (35.7) | 2 (33.3) | - | - |  |  |
| Fluoxetine | 0 (0.0) | 1 (16.7) | - | - |  |  |
| Paroxetine | 1 (7.1) | 0 (0.0) | - | - |  |  |
| Sertraline | 3 (21.4) | 1 (16.7) | - | - |  |  |
| **Delivery type (n vaginal (%))** | 5 (35.7) | 5 (83.3) | 11 (55.0) | 18 (75.0) | 0.281 | 0.99 |
| **Gestational age at birth (weeks, mean (SD))** | 39.1 (1.4) | 39.7 (1.4) | 39.6 (1.3) | 39.5 (1.4) | 0.447 | 0.697 |
| **Gestational diabetes (n, mean (SD))** | 0 (0) | 0 (0) | 0 (0) | 0 (0) | - | - |
| **Infant sex (n male (%))** | 6 (42.9%) | 2 (33.3%) | 11 (55.0%) | 14 (58.3%) | 0.727 | 0.522 |
| **Infant birthweight Z-score (mean (SD))** | -0.01 (1.06) | 0.00 (0.94) | -0.02 (0.62) | -0.05 (0.82) | 0.989 | 0.906 |
| **PlaNET Ancestry<sup>§</sup> (mean (SD))** |  |  |  |  |  |  |
| Coordinate 1 | 0.00 (0.00) | 0.00 (0.00) | 0.01 (0.02) | 0.00 (0.00) | 0.292 | 0.665 |
| Coordinate 2 | 0.07 (0.25) | 0.01 (0.01) | 0.14 (0.32) | 0.12 (0.31) | 0.488 | 0.370 |
| Coordinate 3 | 0.93 (0.25) | 0.99 (0.01) | 0.85 (0.32) | 0.87 (0.31) | 0.460 | 0.369 |

†*p* values are from chi-squared tests for categorical variables and t-tests for continuous variables, \* indicates <0.05, \*\* indicates <0.005.
<sup>§</sup>*p* = 0.59 (chi-squared test) for proportion of SSRI types utilized by SSRI-exposed, maternally-depressed participants versus SSRI-exposed, not maternally-depressed participants.
<sup>§</sup>The PlaNET algorithm outputs DNAm-based ancestry probability values ranging from 0-1, which sum to 1 for each sample. Coordinate 1 is associated with probability of African ancestry, coordinate 2 with East Asian ancestry, and coordinate 3 with European ancestry (46).

**Figure 1.**
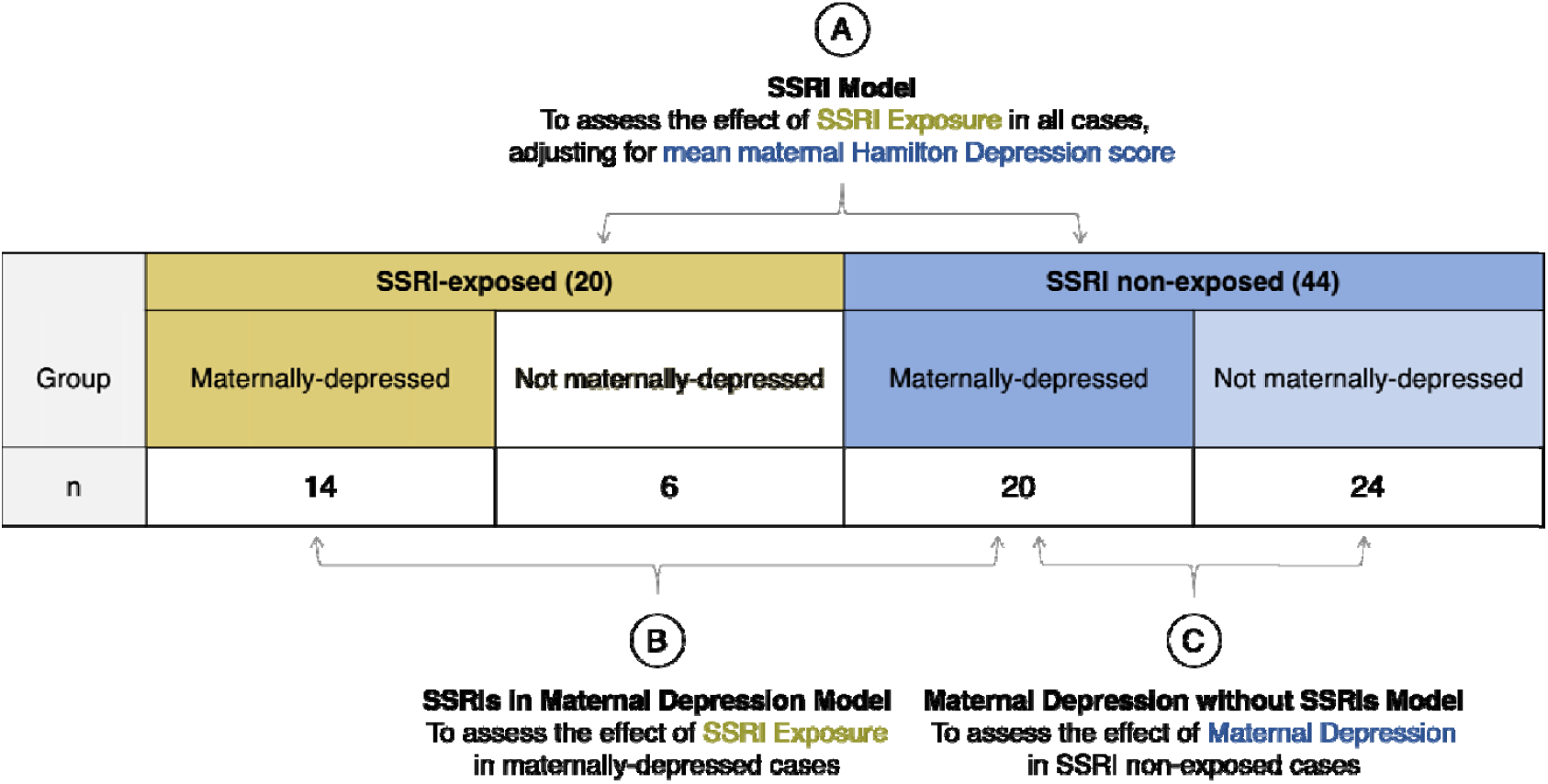
Comparisons for analysis of differential DNAme associated with SSRI exposure and maternal depression in the discovery cohort. **(A)** The SSRI model investigates SSRI exposure in the full cohort (n=64), comparing SSRI-exposed (n=20) to SSRI non-exposed (n=44) cases, adjusting for mean maternal Hamilton Depression score across gestation. **(B)** The SSRIs in maternal depression model investigates SSRI exposure in all maternally-depressed cases (mean maternal Hamilton Depression score >8, n=34), this model compares SSRI-exposed, maternally depressed cases (n=14) to SSRI non-exposed, maternally depressed cases (n=20). **(C)** The maternal depression without SSRIs model investigates the effect of maternal depression in cases not exposed to SSRIs (n=44), this model compares SSRI non-exposed, maternally depressed cases (n=20) to SSRI non-exposed and not maternally depressed cases (n=24).

### Replication cohort

An independent cohort of 34 cases with (i) known SSRI-exposure status and (ii) pre-collected placental DNAme data were obtained from a subset of the Rhode Island Child Healthy Study (RICHS) (44). Cohort participants were treated with one of the following SSRIs: fluoxetine, paroxetine (Paxil), sertraline (Zoloft), citalopram (Celexa), or escitalopram (Lexapro). Assessments of maternal mood were not available for these cases. DNAme data for these 34 cases were previously collected with the Illumina HumanMethylation450 (“450K”) array platform, and are available on the Gene Expression Omnibus (GEO) repository under accession number GSE75248 (44).

### Data processing

Processing and normalization of the Illumina EPIC DNAme data for the discovery cohort is described in full in the Supplementary Methods. First, sample sex (XY chromosome complement) and genetic uniqueness were assessed and confirmed to match the annotated metadata. Next, polymorphic and cross-hybridizing probes were removed from this dataset (47), as were probes with a detection *p* value > 0.01 in > 5% of cases. Subsequently, CpGs with non-variable DNAme in this cohort (n=87,572) were removed from the dataset to decrease multiple test correction burden. Non-variable CpGs were defined as in (48) as CpGs that met both of the following conditions: (i) < 5% range in DNAme beta (β) values between the 10^th^-90^th^ centile, and (ii)previously reported as placenta-non-variable in (49). After processing and dasen + noob normalization (50), 659,036 autosomal CpGs in 64 cases plus 8 pairs of technical replicates remained for analysis. The technical replicates were used to estimate an absolute delta beta (|Δβ|) cut-off for subsequent analyses, based on the average root mean squared error of the technical replicate pairs (0.026) and the average standard error of all CpGs in all cases (0.0048). The highest of these values was selected and rounded up, to establish a |Δβ| cut-off of > 0.03 between groups; anything less than |Δβ| = 0.03 could represent technical noise and was unlikely to be biologically meaningful in this cohort. After processing, the replicate pairs were removed from the cohort and excluded from downstream analysis.

Batch correction was performed after identifying an effect of EPIC array row on DNAme data using Principal Components Analysis, see Supplementary Figure S1. Prior to batch correction, the distribution of cases across EPIC array rows was confirmed to be independent of our primary outcomes of interest: SSRI status (Fisher test p > 0.05) and mean maternal Hamilton depression score (ANOVA p > 0.05), see Supplementary Figure S2. ComBat was used to correct the remaining categorical effect of the “row” variable, with SSRI exposure (yes/no) and mean maternal Hamilton Depression score (continuous) included as variables of interest in the ComBat model matrix as recommended by the *sva* package authors (51). Results for batch-corrected analyses were confirmed in non-batch-corrected data to ensure that false signal was not introduced during batch correction, in accordance with the recommendations outlined in (52), see Supplementary Figure S1.

### Replication cohort

The raw data for the full 335-sample RICHS cohort was downloaded from GEO (GSE75248) and processed analogously to the discovery cohort: first sample sex and genetic uniqueness were checked, data were dasen + noob normalized, and polymorphic and cross-hybridizing probes were removed (47), as were probes with a detection *p* value > 0.01 in > 5% of cases. For a complete description of data processing see the Supplementary Methods. The processed data were subsetted to 34 cases for which gestational SSRI treatment status was recorded in the medical chart. Replication cohort demographics are presented in Table 1.

### Covariate selection

We next sought to identify characteristics that could be associated with SSRI exposure in the discovery and replication cohorts, and that should be included as covariates in statistical models. Demographic variables considered include those presented in Table 1 and Table 2. The discovery cohort was well-balanced across all demographic variables by both SSRI exposure and maternal depression status. Gestational age, infant sex, and PlaNET ancestry were selected for inclusion as additive covariates in linear modelling analyses as it is known that these factors drive large amounts of DNAme variation in the placenta (46,53). The same variables were selected in the replication cohort (infant sex, gestational age at birth, PlaNET ancestry), with the addition of type of delivery (vaginal/other), as delivery type frequency differed by SSRI exposure in the replication cohort only, see Table 1.

Using tools from the *PlaNET* R package (54,55), we also estimated additional variables in the discovery cohort from the DNAme data itself: the proportion of six major placental cell types (cytotrophoblasts, syncytiotrophoblasts, endothelial cells, stromal cells, Hofbauer cells, and nucleated red blood cells), and placental epigenetic age. Cell type proportions were not significantly associated with either SSRI treatment or mean maternal Hamilton Depression score across gestation (t-test for cell type proportions ∼ SSRI exposed (yes/no), all *p* values > 0.05; Pearson correlation for mean maternal Hamilton Depression score and each cell type proportion, all *p* values > 0.05), see Supplementary Figure S3. Accordingly, cell type proportions were not included as covariates in subsequent linear models. Epigenetic age acceleration was calculated as the residual of the Control Placental Clock epigenetic age regressed on gestational age at birth, adjusted for sex, ancestry, and EPIC array row. Intrinsic epigenetic age acceleration (cell-type independent) was calculated similarly, but with additional adjustment for numeric cell type proportions. Placental epigenetic age was not considered as a possible covariate in linear models, but was analyzed separately for associations with SSRI exposure, as previous cord blood studies have reported conflicting relationships between cord blood epigenetic age acceleration and SSRI treatment, so parallel exploration in the placenta was warranted (56,57), see Results.

### Linear modelling to identify placental DNAme associated with SSRI exposure and maternal depression

To identify differential DNAme associated with both SSRI exposure and exposure to maternal depression in the discovery cohort, three linear models were explored, see Figure 1. Linear modelling on M values was conducted using *limma* in R (58,59). In addition to the main effects, all models were adjusted for infant sex, gestational age at birth (in weeks), EPIC array row, and two of three PlaNET ancestry coordinates (46); EPIC array row was included as a covariate according to the recommended implementation of ComBat per the *sva* package authors (51). In (A) the SSRI model, the full cohort was used for a linear model to assess the effect of SSRI exposure (n_exposed_ = 20 versus n_non-exposed_ = 44) on placental DNAme, adjusting for mean maternal Hamilton Depression score across gestation. In (B) the SSRIs in maternal depression model, we used only cases with maternal depression (mean maternal Hamilton Depression score > 8, n=34) and assessed the effect of SSRI exposure in exposed (n=14) versus SSRI non-exposed cases (n=20). In (C) the maternal depression without SSRIs model, we used only cases without SSRI exposure during gestation (n=44), and assessed the effect of maternal depression alone by comparing SSRI non-exposed, maternally-depressed cases (n=20) to not maternally-depressed cases (n=24). Multiple test correction for all models was performed using the Benjamini-Hochberg false discovery rate method (60).

For replication analyses, linear models were run only on the CpGs of interest (M values), identified in the discovery cohort models A-C. Replication linear models were adjusted for infant sex, gestational age at birth, two of three PlaNET ancestry coordinates, and type of delivery (vaginal/other). CpGs were considered to replicate differential DNAme in association with SSRI exposure at a nominal *p* value < 0.05 in the replication cohort.

## RESULTS

### SSRI exposure and maternal depression are not associated with widespread alterations in placental DNAme patterns

First, we assessed in separate models whether DNAme was altered in placentas with SSRI exposure or exposure to maternal depression during gestation. No CpGs were significantly differentially methylated at the commonly used statistical threshold of FDR < 0.05 in any of the three models tested. Plotting FDR against the |Δβ| for all CpG sites tested in the three models demonstrated very few DNAme associations with either SSRI exposure or maternal depression array-wide, see Figure 2. Difference in DNAme (Δβ) is plotted along the X axis and was calculated as Δβ =β_SSRI-exposed_ – β_SSRI non-exposed_. **(B)** Volcano plot for the SSRIs in maternal depression model, differential DNAme associated with SSRI exposure in cases with mean maternal HamD score > 8, (n=34). Difference in DNAme (Δβ) is plotted along the X axis and was calculated as Δβ = β_SSRI-exposed_ – β_SSRI non-exposed_. **(C)** Volcano plot for the maternal depression without SSRIs model, differential DNAme associated HamD score > 8 across gestation, in SSRI non-exposed cases, (n=44). Difference in DNAme (Δβ) is plotted along the X axis and was calculated as Δβ = β_Hamilton > 8_ – βHamilton ≤ 8.

**Figure 2.**
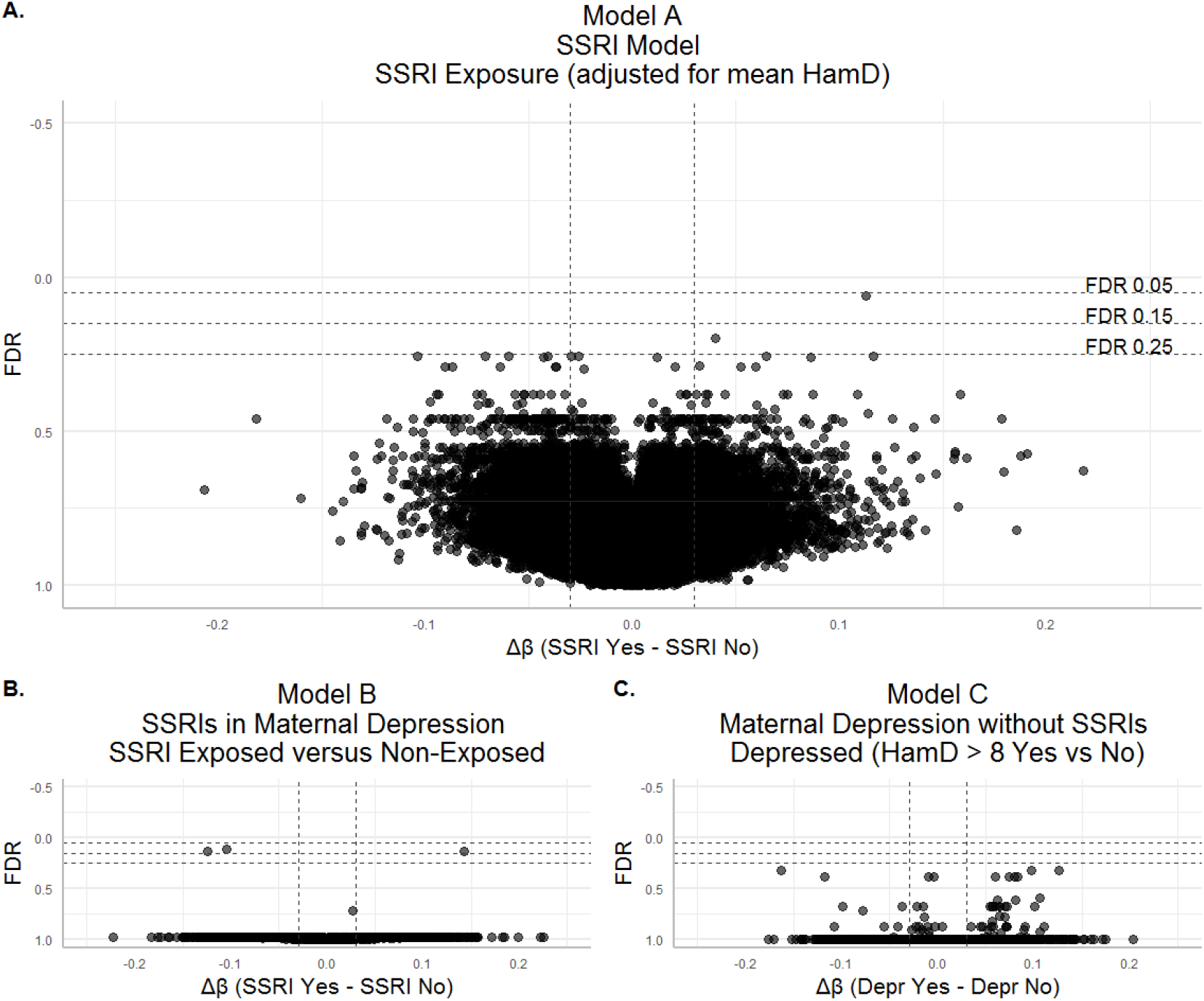
Volcano plots for differential DNAme in association with SSRI exposure and categorical depression. For all plots HamD refers to the mean maternal Hamilton Depression score across gestation for each individual. False discovery rate (FDR) is shown along the Y axis with more significant (lower FDR) values at the top of the plot. Vertical dashed intercepts demarcate |Δβ| = 0.03, horizontal dashed intercepts indicate from top to bottom FDR = 0.05, FDR = 0.15, and FDR = 0.25 **(A)** Volcano plot for the SSRI model, investigating differential DNAme associated with SSRI exposure adjusted for mean maternal HamD score, (n=64).

Since the relatively small sample size of the discovery cohort could limit our ability to detect significant between-group DNAme differences, in addition to the standard threshold of FDR < 0.05, we also evaluated CpGs that met more relaxed thresholds of FDR < 0.15 and FDR < 0.25. As these thresholds are associated with increased expected proportions of false positive findings, we have labelled FDR < 0.15 hits “moderate-confidence” and FDR < 0.25 “low-confidence”. For both FDR < 0.15 and FDR < 0.25 thresholds, we maintained a minimum effect size threshold of |Δβ| > 0.03.

At these relaxed thresholds, in the SSRI model (model A) adjusted for mean maternal Hamilton Depression score, one CpG was differentially methylated at FDR < 0.15, and one more at FDR < 0.25, see Figure 2 and Supplementary Table S1. The one differentially methylated CpG at FDR < 0.15 (cg12900404) exhibited a Δβ of +0.11 (higher average DNAme β value in SSRI-exposed placentas). Loosening the threshold to FDR < 0.25, one additional CpG was differentially methylated by SSRI exposure status (cg20877313), and was also more highly methylated in SSRI-exposed placentas (Δβ=+0.04). In the SSRIs in maternal depression model (model B), three CpGs were differentially methylated at the moderate-confidence threshold of FDR < 0.15 (cg12655501, cg26993610, cg14340829). These CpGs all had an average difference in DNAme between SSRI-exposed and SSRI non-exposed cases of |Δβ| > 0.10. In the maternal depression without SSRIs model (model C), no CpGs showed evidence of maternal depression-associated differential DNAme at any FDR threshold. For a description of all CpGs that satisfied the moderate and low-confidence FDR thresholds from the SSRI models (A, B) and SSRIs in maternal depression model (C), including test statistics and overlapping genes, see Table 3. Summary statistics of linear modelling from all CpGs tested are presented in Supplementary Tables S2-S4. Boxplots showing average DNAme β values in SSRI-exposed versus SSRI non-exposed cases for the differentially methylated CpGs are shown in Figure 3.

**Table 3.** Top CpGs with differential DNAme by SSRI-exposure status. Linear modelling p value and false discovery rates (FDR) shown, Δβ is difference in DNAme calculated as Δβ = β_SSRI-exposed_ – β_SSRI non-exposed_. hg19 chromosome (Chr) and coordinates (Position) for each CpG are indicated, as are genes that each CpG overlaps, and whether each CpG is covered by a probe on the 450K DNA methylation array. A and B in the Model Column refer to model A and model B, DMR refers to the differentially-methylated region analysis.

| Model | CpG | $p$<br>value | FDR | FDR<br>threshold | $\Delta\beta$ | Chr:Position | Gene<br>Symbol | Relation<br>to Gene | 450K<br>locus? |
| --- | --- | --- | --- | --- | --- | --- | --- | --- | --- |
| A | cg12900404 | 9.11e-08 | 0.06 | Moderate<br>confidence | 0.11 | 2:225811669 | *DOCK10* | 1<sup>st</sup> Exon | - |
| A | cg20877313 | 6.05e-07 | 0.20 | Low<br>confidence | 0.04 | 12:56881753 | *GLS2* | 1<sup>st</sup> Exon | Yes |
| B | cg12655501 | 6.07e-07 | 0.14 | Low<br>confidence | -<br>0.12 | 1: 115603524 | *TSPAN2* | Body | - |
| B | cg26993610 | 6.26e-07 | 0.14 | Low<br>confidence | 0.14 | 1:115605466 | *TSPAN2* | Body | - |
| B | cg14340829 | 1.80e-07 | 0.12 | Low<br>confidence | -<br>0.10 | 8:923973 | - | - | Yes |
| DMR | cg14921691,<br>cg06762403 | - | 1.95e-29 | - | 0.06 | 12:<br>56325797-56325867 | *DGKA* | 5'UTR,<br>1<sup>st</sup> Exon | Yes |

**Figure 3.**
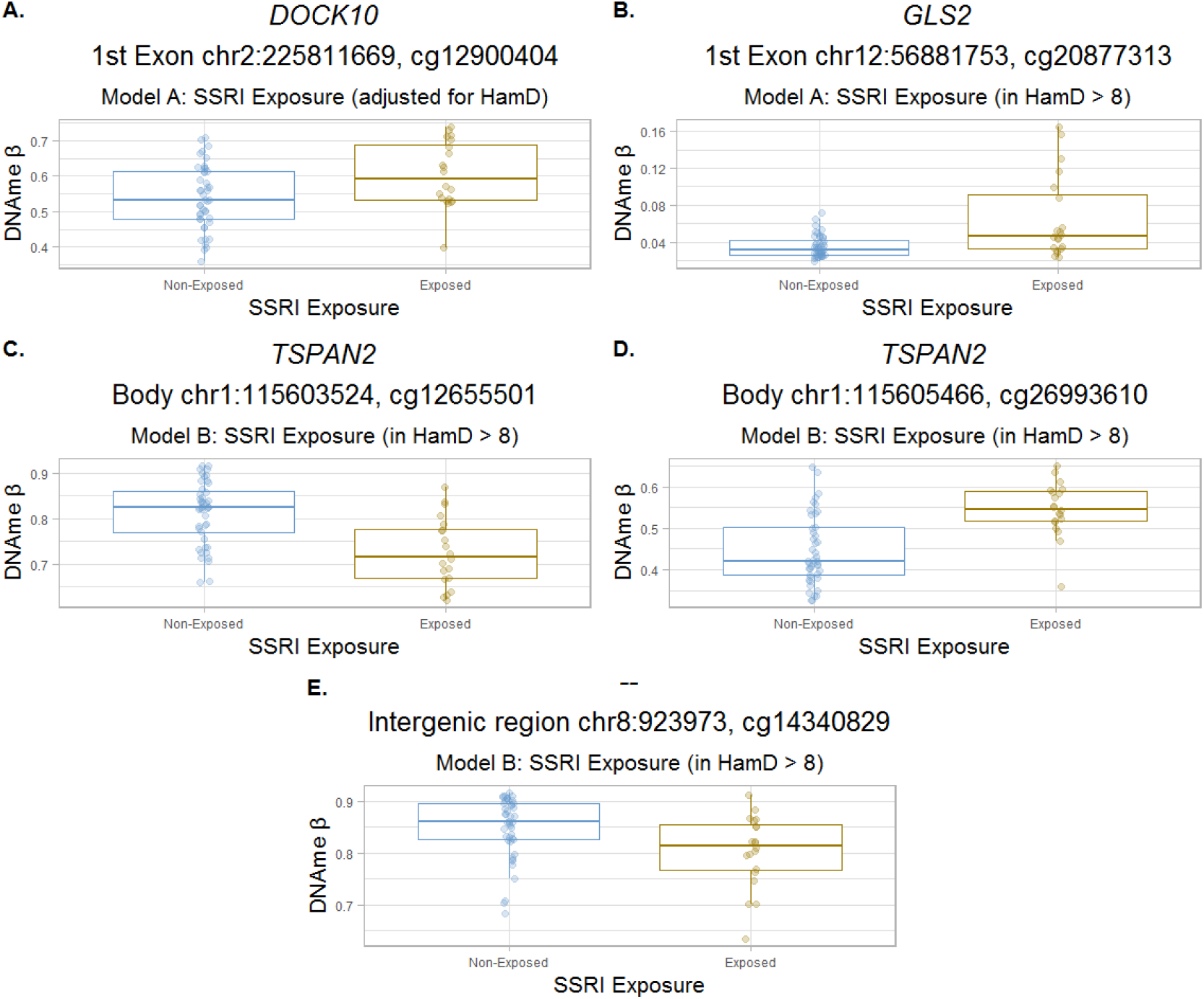
Boxplots of differentially methylated CpGs by SSRI exposure. For all plots, points are colored by SSRI exposure (blue = SSRI non-exposed, dark yellow = SSRI-exposed), and boxplots indicate mean DNAme β value ± one standard deviation. **(A)** Boxplot of cg12900404, identified in the SSRI model, model A. **(B)** Boxplot of cg20877313, identified in the SSRI model, model A. **(C)** Boxplot of cg12655501, identified in the SSRIs in maternal depression model, model B. **(D)** Boxplot of cg26993610, identified in the SSRIs in maternal depression model, model B. **(E)** Boxplot of cg14340829, identified in the SSRIs in maternal depression model, model B.

A differentially methylated region (DMR) analysis was also conducted, to increase power for discovering DNAme-associations by grouping signatures from multiple neighboring CpG sites to reduce the number of statistical comparisons. DMR analysis was only conducted for the SSRI model to enable utilization of the entire cohort of cases and maximize statistical power.

M values were assessed for being part of DMRs for which average DNAme differed by SSRI exposure status using the *DMRcate* R package (FDR < 0.25, lambda = 1000, C = 2) (61). Only one DMR was identified with an average DNAme β value that differed by SSRI exposure, this DMR comprised of two CpGs 70 base pairs apart (cg06762403 and cg14921691) on chromosome 12, overlapping the *DGKA* gene. The average Δβ across the region was +0.06, indicating higher DNAme in SSRI-exposed placentas, see Table 3 and Supplementary Figure S4.

### Few placental DNAme differences associated with SSRI exposure in sex-stratified models

As the effects of SSRI exposure might differ by sex, we next sought to assess whether unique SSRI-exposure DNAme associations arose in placentas of male versus female infants. Sex-stratified linear models were run in males (n=33, 24% SSRI-exposed) and females (n=31, 39% SSRI-exposed) to test for SSRI-exposure associated DNAme, adjusting for mean maternal Hamilton Depression Score, gestational age at birth, EPIC array row, and PlaNET ancestry. At an FDR < 0.25 and |Δβ| > 0.03, no CpGs were differentially methylated by SSRI exposure in males. In females, at an FDR < 0.25 and |Δβ| > 0.03, three CpGs (cg15849349, cg07481545, and cg03905236) were differentially methylated in association with SSRI exposure (Supplementary Figure S5).

We previously identified 162 CpGs with robust sex differences in placental DNAme patterns that replicated in an independent dataset (62). To investigate sex in another context, we tested whether these previously-ascertained sex-differentially-methylated CpGs were also differentially methylated by SSRI exposure status. All 162 replicated CpGs were considered, and were subjected to linear modelling in the discovery cohort for SSRI exposure, adjusting for mean maternal Hamilton Depression Score, gestational age at birth, EPIC array row, and PlaNET ancestry. No CpGs met significance at FDR < 0.05. At the low-confidence FDR < 0.25 threshold, three CpGs (cg22515303, cg27003571, cg26136722) had lower DNAme in SSRI-exposed placentas than non-exposed placentas, in addition to the previously identified sex effect at these loci (Supplementary Figure S6).

### No replication of DNAme signature associated with SSRI exposure in an independent dataset

We next intended to assess the reproducibility of the five CpGs with SSRI-associated differential DNAme identified in the SSRI model and SSRIs in maternal depression model (Table 3), and the two DMR CpGs. As the replication cohort was run on the Illumina 450K array, only four of these seven discovery cohort CpGs could be assessed for replication (cg14340829, cg20877313, cg06762403, cg14921691). A linear model was applied to the M values from these four CpGs in the replication cohort to test for differential DNAme associated with SSRI exposure, adjusting for infant sex, gestational age at birth, mode of delivery, and PlaNET ancestry. None of the four CpGs replicated differential methylation at a nominal *p* value < 0.05 in the replication dataset, see Supplementary Figure S7.

### Placental epigenetic age acceleration not associated with SSRI exposure or maternal depression

Epigenetic age is a useful dimension-reduction technique to analyze relationships between global DNAme patterns and a variety of health outcomes. Altered epigenetic age relative to chronological age (“epigenetic age acceleration”) has been associated with several negative health outcomes including overall greater burden of disease and higher rates of all-cause mortality (63,64). Specific to depression, major depressive disorder is associated with increased epigenetic age acceleration in adult blood (65). Umbilical cord blood epigenetic age deceleration was initially reported in maternal depression (56), but interestingly this association has since been demonstrated to be largely related to the confounding signature of maternal SSRI treatment rather than depression itself (57).

Using a recently published placental epigenetic clock (55), we calculated epigenetic age acceleration in the discovery cohort. We then tested for associations between epigenetic age acceleration metrics and either SSRI exposure or mean maternal Hamilton Depression score using linear models. Neither epigenetic age acceleration nor intrinsic epigenetic age acceleration (cell-type independent) were significantly associated with either SSRI exposure or maternal depression, see Supplementary Figure S8.

### Replication of previously reported differential methylation candidates associated with SSRI-exposure and maternal depression in the discovery cohort

A previous study identified 16 CpGs with altered placental DNAme associated with maternal depression (treated as a categorical variable, where depression was maternal EPDS > 10) at FDR < 0.05 (no |Δβ| threshold) (66). EPDS measurements were also collected for the discovery cohort cases in our study, and were significantly collinear with mean maternal Hamilton Depression scores across gestation, see Supplementary Figure S9. To assess whether these previously-reported CpGs were also differentially methylated in association with maternal EPDS in the discovery cohort, we ran a linear model on the DNAme M values from these 16 CpGs, adjusting for infant sex, gestational age at birth, ancestry, and EPIC array row. One CpG (cg06670742) satisfied a nominal *p* value < 0.05 in our cohort, and had higher DNAme in cases with higher mean maternal EPDS scores (FDR < 0.05, Δβ=+0.003). Thus, differential DNAme of this CpG did replicate in our study, however the effect size in our cohort was very small and does not exceed the technical threshold we set of |Δβ| > 0.03 (Figure 4).

**Figure 4.**
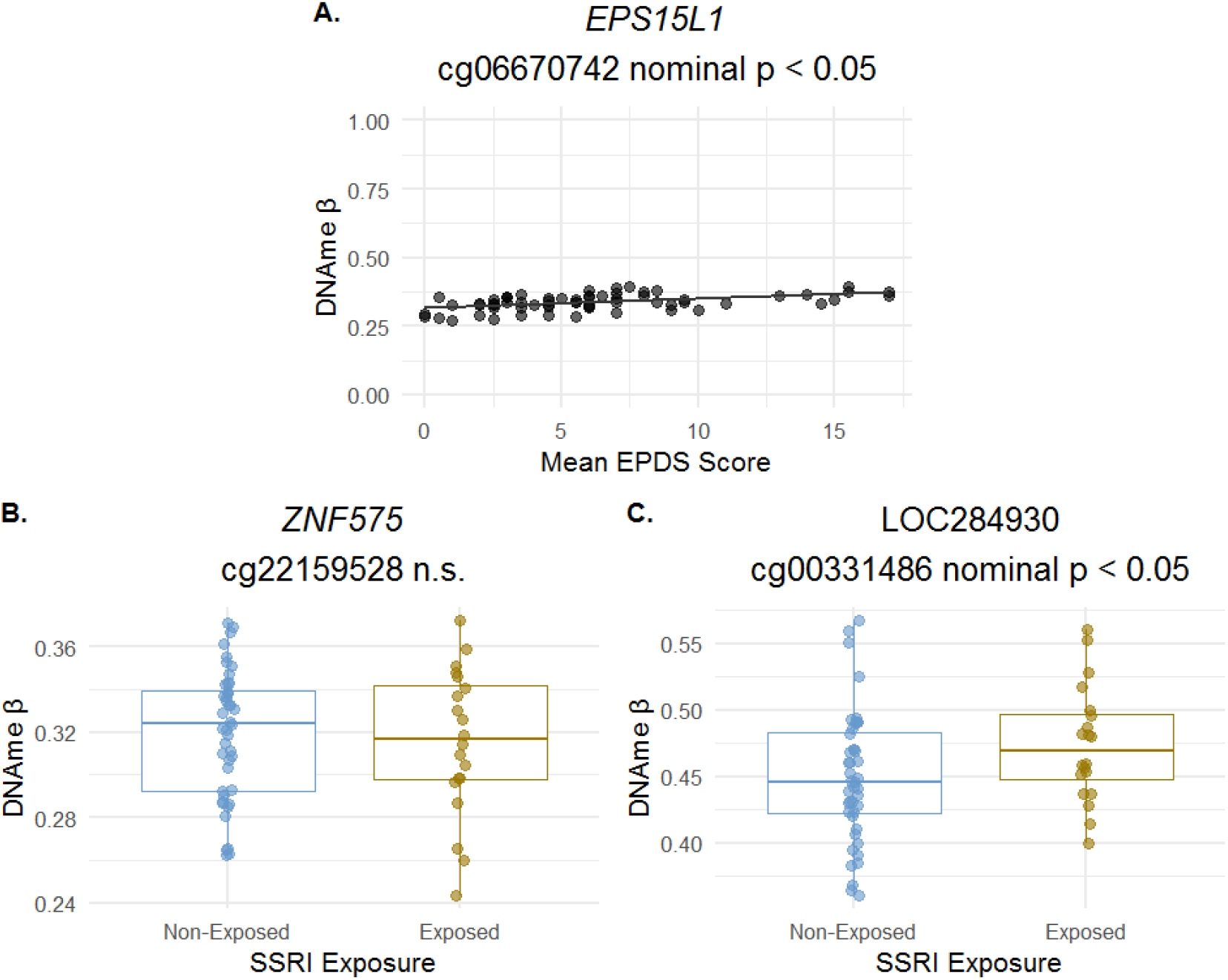
Assessment of literature candidates for SSRI and depression-associated DNAme in the discovery cohort. **(A)** Scatterplot showing the association between DNAme and maternal Edinburgh Postnatal Depression Score (EPS) at cg06670742 which was previously found to b e differentially methylated in association with maternal EPDS score in Tesfaye et al. (2021) (37) and is differentially methylated by SSRI exposure at FDR < 0.05 in our discovery cohort. DNAme β value at this CpG is plotted along the Y axis, mean maternal EPDS score is plotted along the X axis and each point is a case; Δβ per unit increase in EPDS is Δβ=+0.003. **(B)** Boxplot showing mean DNAme β value ± one standard deviation by SSRI exposure status at cg22159628 in ZNF575, previously found to be differentially methylated in cord blood with antidepressant exposure. Linear modelling p value = 0.67, points are colored by SSRI exposure (blue = SSRI non-exposed, dark yellow = SSRI-exposed), n.s. = not significant. (C) Boxplot showing mean DNAme β value ± one standard deviation by SSRI exposure status at cg00331486 in LOC284930 on chromosome 22, previously found to be differentially methylated in cord blood by SSRI exposure status.

Three previous studies investigated genome-wide DNAme alterations associated with SSRI treatment in a prenatal tissue (cord blood). An early study by Non et al. (2014) found no association between SSRI exposure and cord blood DNAme at any CpG loci (67). Cardenas *et al*. (2019) reported one differentially methylated CpG in the gene body of *ZNF575* associated with maternal antidepressant treatment in a cohort of cord blood samples (68). In a different cohort, Kallak et al. (2021) reported 13 CpGs differentially methylated by SSRI exposure status in cord blood (69). Though cord blood DNAme is very distinct from placenta, we sought to evaluate whether any of the 14 CpGs reported by Cardenas et al. (68) or Kallak et al. (69) were also differentially methylated by SSRI exposure status in the placenta. At the CpG reported by Cardenas et al. (cg22159528) we found no difference in DNAme by SSRI exposure (nominal *p* value > 0.05), see Figure 4. At one of the 13 CpGs reported by Kallak et al., we found nominally significant (*p* < 0.05) differential DNAme associated with SSRI exposure status in the placenta in the same direction as the original cord blood association: higher DNAme in SSRI-exposed cases. This was cg00331486 in LOC284930 on chromosome 22, which encodes a MYC-interacting long noncoding RNA species (69).

## DISCUSSION

In this study, we examined genome-wide placental DNAme patterns associated with SSRI exposure or maternal depression during gestation. Taken together, our results suggest the absence of a major impact of SSRI antidepressant exposure on the placental DNA methylome. Specifically, we found no differentially methylated CpGs associated with either SSRI exposure or maternal depression that met the standard significance threshold of FDR < 0.05. These results hold despite known alterations elicited by SSRIs to maternal gestational serotonin signalling and infant outcomes, suggesting that the effects of SSRI exposure on infant outcomes are not associated with a coincident DNAme signature in the placenta.

At more relaxed statistical thresholds, differential DNAme analysis highlighted five CpGs associated with SSRI exposure status. Three of the five differentially methylated CpGs overlapped the following genes, respectively: *DOCK10* (CpG not present in replication cohort), *GLS2* (CpG did not replicate), and *TSPAN2* (CpG not present in replication cohort). The *DOCK10* gene encodes the Dopamine Receptor Interacting Protein (70), and was recently found to be one of the twelve most predictive mRNA biomarkers of depression in a gene expression study of adult whole blood (71). These researchers identified that *DOCK10* expression was higher in association with a more positive, less depressed mood (71). *DOCK10* is also a target of increased expression by the action of the antidepressant ketamine(71). *TSPAN2* encodes Tetraspanin-2, which mediates signal transduction in processes related to cellular growth and development (72). *Tspan2* has been reported to be upregulated in the rat hippocampus after chronic exposure to the SSRI fluoxetine (73). We also identified one differentially methylated region in the 5’ untranslated region of *DGKA*, with higher DNAme in SSRI exposed placentas. *DGKA* encodes a diacylglycerol kinase that is involved in intracellular signalling (74), and *DGKA* activity was recently reported to be inhibited by Ritanserin, a pharmaceutical serotonin receptor type 2 (5-HTR2) antagonist that is not currently in clinical use (75).

Inherent to studying the impact of SSRI exposure are the concurrent, and potentially confounding, effects of maternal mood disturbances. As such, studies must be carefully designed to interrogate the effects of either SSRI treatment or depression exposure while accounting for the other factor. Only a limited placental DNAme signature has so far been associated with maternal mood disorders (66), and no work has been conducted to our knowledge interrogating genome-wide placental DNAme signatures associated with SSRI exposure. Tesfaye *et al*. (66) investigated placental DNAme in association with maternal depression, though SSRI treatment or antidepressant use was not reported on or investigated and could underlie some of these associations. Of the 16 CpGs they identified as differentially methylated by maternal depression status across six gestational time points, only one CpG in the *EPS15L1* gene was also differentially methylated our study by mean maternal EPDS score across gestation (FDR < 0.05), and although the effect was in the same direction, the effect size (Δβ = +0.003 with each unit increase in mean EPDS) was well below our biological |Δβ| threshold of > 0.03, and therefore may not be a biologically meaningful change in DNAme.

In non-placental tissue, Kallak et al. (69) identified 13 CpGs that were differentially methylated in cord blood in association with SSRI exposure, one of which replicated at nominal significance in the same direction (higher DNAme in SSRI-exposed cases) in our cohort. Cardenas et al. also identified one CpG in the gene body of *ZNF575* (cg22159528) with lower DNAme in antidepressant-exposed cord blood samples from Project Viva, which they replicated in an external cohort (Generation R Study) (68). In our cohort, placental DNAme at this CpG was not associated with SSRI exposure. Although cord blood and placenta are both conceptus-derived tissues relevant to prenatal development and early life, they have unique origins (embryonic versus extraembryonic lineages, respectively) and thus distinct DNAme profiles are expected. Our finding that cord blood differential DNAme does not replicate at high rates in the placenta may suggest a cord-blood-specific DNAme signature at these CpGs in association with maternal SSRI treatment.

In sex-stratified analyses, three CpGs were differentially methylated by SSRI exposure in females: two in intergenic regions on chromosomes 4 and 6 respectively, and one in the 5’ untranslated region of the *SH3GL3* gene. *SH3GL3* encodes the Endophilin-A3 protein, which interacts selectively with the Huntingtin protein to promote the formation of polyglutamine-containing protein aggregates (76). Promoter DNA methylation of the *SH3GL3* gene was previously inversely correlated with its expression in human colon cancer cell lines versus matched double knockout cell lines for DNMT1 and DNMT3b (77). Lower DNAme upstream of the promoter of the *SH3GL3* gene in SSRI-exposed female placentas may indicate higher expression of this gene relative to the placentas from SSRI non-exposed females, which should be followed up in future work. In a second sex-focused analysis we tested whether previously reported sex-biased CpGs (62) were differentially methylated by SSRI status, three CpGs met the low-confidence FDR < 0.25 threshold (two in the 5’ untranslated region of *GTDC1*, one in the gene body of *C14orf132*). Together, these analyses suggest that there may be a sex-biased signature of SSRI treatment on the placental DNA methylome or transcriptome, and future studies in this field should be designed to analyze sex as a variable of interest.

The major strength of our study lies in the curation of both SSRI treatment and maternal depression information per participant. This prospective design enabled us to investigate the potential confounding effects of these two variables in controlled contexts (i.e. studying SSRI exposure while adjusting for depression scores, and studying the effect of maternal depression in SSRI non-exposed samples). Additionally, in this study we measured genome-wide DNAme in an effort to reduce bias that arises from candidate gene or CpG pyrosequencing studies, and utilized the most modern Illumina EPIC array platform to capture the highest resolution data currently afforded by a DNAme microarray. Further strengths of our study include strict effect size and significance thresholds, the presence of an external replication cohort, the ability to account for covariates such as genetic ancestry and cell type proportions, and the ability to stratify analyses by sex.

Limitations of this work include SSRI exposure being defined as maternal treatment with any of six different SSRIs during gestation. It is possible that the SSRI subtypes have different effects on placental DNAme, though this study was not able to undertake sub analysis by SSRI type, and future work should consider this. The extent to which SSRI treatment improves maternal depression scores (i.e. remittance of symptoms) may also be associated with placental DNAme patterns and should be investigated in future longitudinal cohorts. Lastly, although sample size was relatively small, our cohort is among one of a handful of prospectively recruited cohorts that we are aware of with placental tissue collected alongside maternal depression and SSRI treatment information, including the BASIC and Generation R Studies (78,79). Cord blood DNAme data exists for both of these cohorts and has previously been studied in association with maternal SSRI exposure (68,69). Cohorts such as these, with extensive clinical records of maternal mood and medication use across gestation are incredibly valuable resources to investigate how depression status and antidepressant medications may interact with fetal outcomes.

In conclusion, our work demonstrates that limited placental DNAme alterations are associated with maternal SSRI exposure; this work agrees with earlier findings of a similarly limited placental DNAme signature of maternal depression. Future work in larger cohorts will determine whether other molecular alterations may co-exist with maternal mood disorders or SSRI treatment, such as altered placental gene expression patterns. As larger cohorts with clinical characterization of maternal SSRI treatment and prenatal depressed mood become available, genetic sequence variation, gene expression patterns, and DNA methylation alterations should all be considered, as well as their interactions.

## Data Availability

All data produced are available online under the Gene Expression Omnibus (GEO) accession number GSE203396.

## DECLARATIONS

### Data availability statement

The raw and preprocessed EPIC array data and corresponding sample metadata supporting the conclusions of this article are available on the Gene Expression Omnibus (GEO), under the accession number <u>GSE203396</u>.

### Ethics approval and consent to participate

Participants were recruited as part of a prospective, longitudinal cohort approved by the University of British Columbia Children’s and Women’s Health Centre of British Columbia Research Ethics Board (UBC C&W REB) (H12-00733 (40)), this sub study was also approved by the same ethics boards (H16-02280).

## Authors’ contributions

AMI conducted the analyses and wrote the manuscript, and contributed to sample processing, quality control, and testing. CK conducted a preliminary analysis of the effect of SSRI exposure on an earlier version of this data, and contributed to sample processing, quality control, and testing. MSP is the NIH project manager, curated and organized clinical data, contributed to sample processing, quality control, and testing, and manuscript editing. UB conducted clinical data collection. AK processed and normalized the discovery cohort EPIC array data. EMP and JMS were involved in data management, sample metadata collation and establishment of the RedCap database, and participated in randomization. ÉPC oversaw project administration and data curation of this cohort. AB performed data curation for the RICHS cohort cases, CJM is cohort owner of the RICHS data and organized data curation for this set of cases. CV contributed to the conceptualization and design of this project, and also contributed to funding acquisition.

TFO and WPR were responsible for project administration, funding acquisition, and supervision of the SSRI cohort recruitment and the DNAme data generation aims of the project, respectively, and were major contributors in writing and revising the manuscript. TO designed the original study. AMI, CK, MSP, UB, AK, EMP, JMS, ÉPC, CV, TFO, and WPR all participated in experimental design. All authors read, revised, and approved the final version of the manuscript.

## Acknowledgements

We acknowledge all cohort owners and collaborators on the “Using ‘omics to build an atlas of placental development and function across pregnancy” (NIH Project). Thank you to Dr. Michael S. Kobor and the BCCHRI Microarray Core Facilities for support with Illumina EPIC DNAme runs. We thank Robinson lab members Ruby Jiang, Giulia F. Del Gobbo, and Desmond Hui for their work in sample processing, quality control, and testing, thank you to Victor Yuan and Ziqi Liu for pilot data processing for cases from this cohort. Finally, we sincerely thank the pregnant individuals and families that contributed placental samples to this study.

