## Supplementary Methods, Figures, & Tables for "Profiling placental DNA methylation associated with maternal SSRI treatment during pregnancy" (from another version of this preprint: version 1, 2022-06-22)

### Supplementary Information

#### Supplementary Methods

##### *Discovery cohort*

Placental chorionic villi samples were obtained for each of the 64 cases in the discovery cohort. As our previous work has shown that DNA methylation (DNAme) patterns vary between placental sampling sites [1, 2], to obtain a representative sample of the whole placenta four independent villi biopsies (1.5-2 $cm^3$ sites from different cotyledons) were taken from the fetal side of each placenta. DNA was extracted from each of the four villi sites using a standard salting-out procedure [3]. Extracted DNA from each of the four sites per placenta was then combined in equimolar amounts for DNAme analysis.

Combined placental DNA from the 64 placental samples was purified using the Qiagen DNeasy Blood and Tissue kit (Qiagen, CA, USA), and 750 ng of DNA per placenta was bisulfite converted using the Zymo EZ DNA Methylation kit (Zymo Research, USA). Bisulfite-converted DNA was whole-genome amplified, enzymatically fragmented, and hybridized to the array per Illumina's protocol for the Infinium MethylationEPIC BeadChip [4]. The cases from the discovery cohort were assayed as part of a larger 424-sample placental DNAme cohort, with all samples similarly extracted and pooled from four placental sites, as well as purified and bisulfite-converted before loading on the array chips.

To minimize technical effects of sample processing, the 64 cases from the discovery cohort were run within the same EPIC array batch distributed across 9 EPIC chips. The discovery cohort cases were carefully randomized across the chips and the chip rows (1-8) by SSRI exposure (yes/no) and infant sex assigned at birth (male/female) and were well-distributed by mean maternal Hamilton Depression score (Supplementary Figure 2). The remaining cases from the 424-sample cohort were run in subsequent array batches.

Raw IDAT files from all 424 samples were read directly into R (version 4.1.1) using the *minfi* and *ewastools* packages [5–8]. To exclude the possibility of sample mislabelling, contamination, or misloading, genetic uniqueness of all samples was confirmed by clustering on the 59 explicit genotyping ("rs") probes present on the EPIC array. Placental sex chromosome complement was confirmed to match reported infant sex at birth using the *ewastools*::*check_sex()* function [6]. Poor quality probes from the following categories were removed from the dataset: detection $p$ value > 0.01 or beadcount < 3 in > 5% of samples (n=4,351); cross-hybridizing probes and probes with single nucleotide polymorphisms within 5 base pairs of the CpG (n=99,360) [9]; and probes located on the X or Y chromosome (n= 19,637). After sample quality control, probe filtering, and dasen+noob normalization [10], 746,608 autosomal CpGs remained; the 64 discovery cohort samples were then extracted from the 424-sample cohort for separate analysis of SSRI exposure. As a final step prior to analysis, CpGs with non-variable DNAme in the 64 sample discovery cohort samples were removed from the dataset (n=87,572); non-variable DNAme was defined as in [11]: probes with less than a 5% range in DNAme $\beta$ values between the $10^{th}$ – $90^{th}$ centile that overlapped with CpGs previously reported to be placenta-non-variable in [12]. The final dataset subjected to analysis comprised of 659,036 CpGs in 64 samples.

*Replication cohort*

Raw IDAT files for 335 samples from GSE75248 were read into R (version 4.1.1) using the minfi and ewastools packages [5–8]. Genetic uniqueness and sample sex were confirmed to match reported metadata, following the same methods used in the discovery cohort. Poor quality probes were removed from the dataset using the same criteria as applied in the discovery cohort and included: probes with detection *p* value > 0.01 or beadcount > 3 in > 5% of samples (n=4,312); cross-hybridizing probes and probes with single nucleotide polymorphisms within 5 base pairs of the CpG (n=60,460) [9]; probes located on the X or Y chromosome (n=11,648). After sample quality control and probe filtering, dasen+noob normalization was applied [10], and 411,896 autosomal CpGs remained for analysis. The data were then filtered to only the 34 samples for which gestational SSRI treatment information was available. For replication of differentially methylated CpGs in the discovery cohort, the CpGs to be assessed were selected from the replication cohort dataset, and a linear model was run only on these loci to assess replication of differential DNAme associated with SSRI exposure. This linear model was adjusted for sex, gestational age at birth, PlaNET ancestry, and mode of delivery (vaginal versus Caesarean section); mode of delivery was included as an adjustment covariate as frequency of vaginal delivery differed in association with SSRI exposure in the replication cohort (chi-squared *p* value 0.016), see main manuscript Table 1.

#### Supplementary Figures

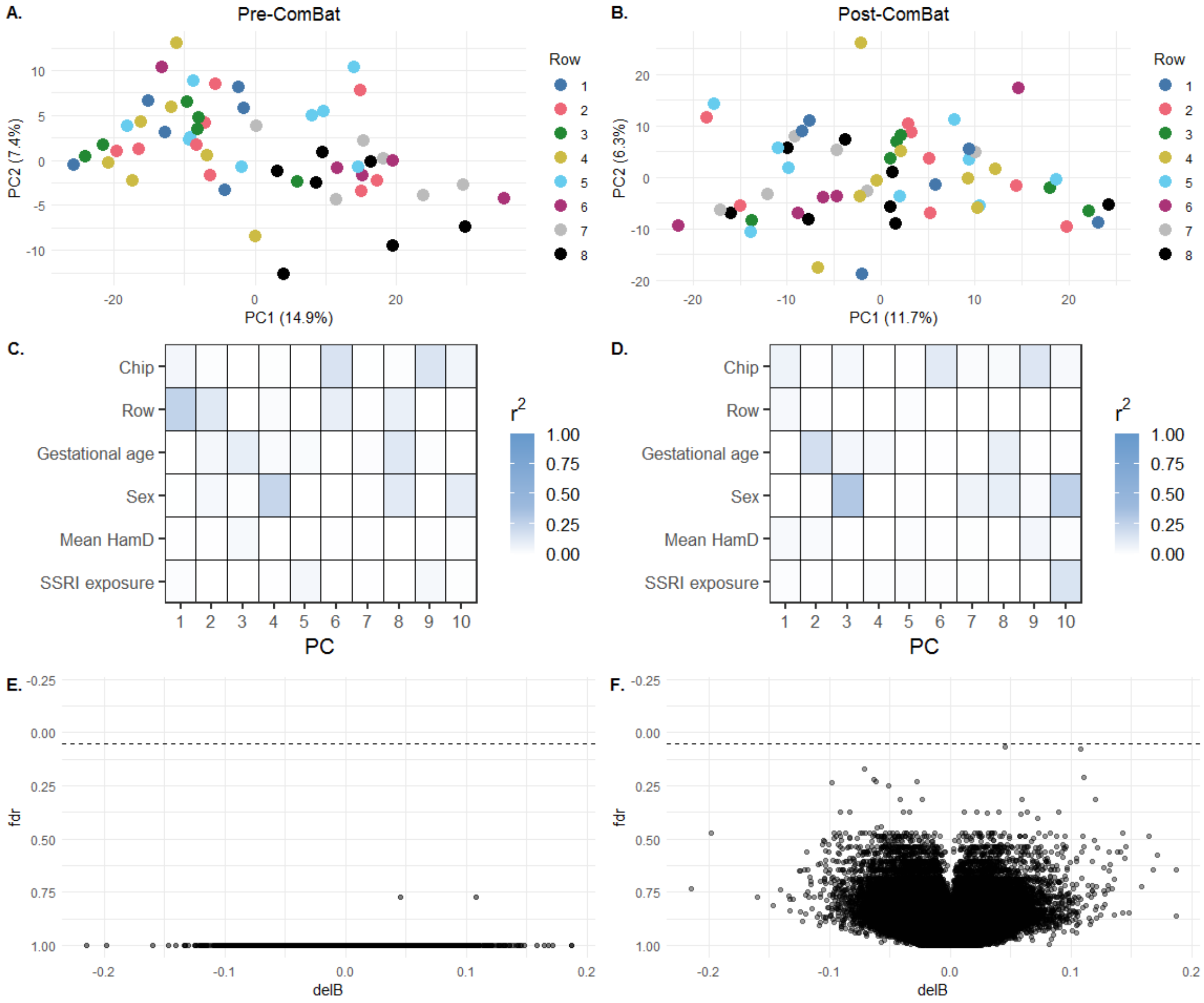


**Supplementary Figure 1. Row effect before and after batch correction with ComBat.** Pre- and post-ComBat batch correction. **(A)** Scatterplot of PC1 versus PC2 before ComBat colored by row, **(B)** scatterplot of PC1 versus PC2 after ComBat for row effect, each point represents a sample, points are colored by row in which the sample was placed on the EPIC array chips. **(C)** PCA heatmap [13] showing strength of association (linear model R squared) between PC scores and sample metadata variables, showing row effect associated with PC1, **(D)** PCA heatmap showing strength of association (linear model R squared) between PC scores and sample metadata variables after ComBat row effect correction. **(E)** Volcano plot of data testing for SSRI exposure in processed and normalized data, horizontal intercept shows FDR = 0.05. **(F)** Volcano plot in ComBat-adjusted data testing for DNAme alterations in association with SSRI exposure, horizontal intercept shows FDR = 0.05.

A.

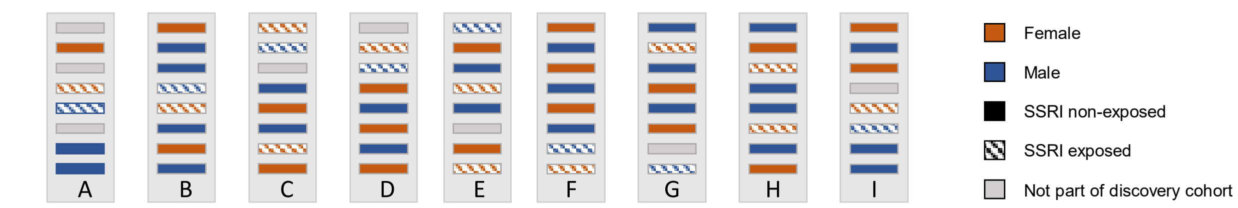


B.

| | SSRI exposed | SSRI non-exposed | p value* |
|---|---|---|---|
| n | 20 | 44 | |
| Row (n) | | | |
| 1 | 2 | 5 | 0.99 |
| 2 | 3 | 6 | |
| 3 | 2 | 5 | |
| 4 | 3 | 5 | |
| 5 | 3 | 6 | |
| 6 | 2 | 5 | |
| 7 | 2 | 6 | |
| 8 | 3 | 6 | |

*p values are from chi-squared tests for categorical variables and t-tests for continuous variables

C.

| Row | n | Hamilton Depression Score (mean (SD)) | p value* |
|---|---|---|---|
| 1 | 7 | 8.07 (3.80) | 0.097 |
| 2 | 9 | 10.28 (5.41) | |
| 3 | 7 | 8.43 (2.30) | |
| 4 | 8 | 9.38 (4.74) | |
| 5 | 9 | 8.06 (3.86) | |
| 6 | 7 | 5.43 (1.97) | |
| 7 | 8 | 11.25 (3.64) | |
| 8 | 9 | 6.67 (3.50) | |

**Supplementary Figure 2. Randomization of samples across EPIC array chips and rows. (A)** Sample distribution across the nine Illumina EPIC array chips A-I. Sample position among the chips (A-I) and chip rows (1-8) was randomized for SSRI exposure (hashed indicates SSRI exposed, solid fill indicates non-exposed) and offspring sex assigned at birth (orange indicates female, blue indicates male). **(B)** Table of counts of sample SSRI exposure status across chip rows. **(C)** Table of mean sample Hamilton Depression scores across chip rows.

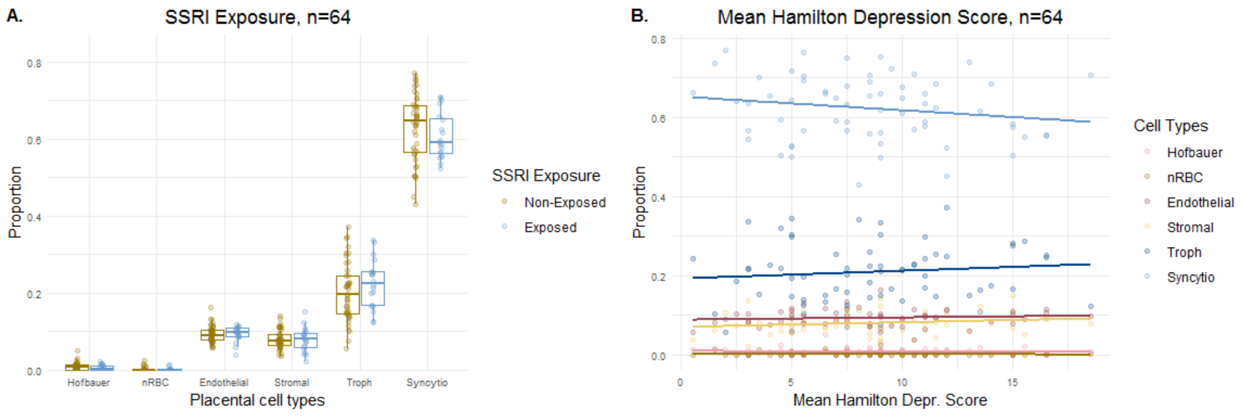


**Supplementary Figure 3. Placental cell type proportions do not vary with SSRI exposure or mean maternal Hamilton depression score. (A)** Estimated proportions of six major placental cell types are not associated with (A) SSRI exposure. Data points are colored by SSRI exposure status (blue = SSRI non-exposed, dark yellow = SSRI-exposed). **(B)** Estimated cell type proportions are not associated with mean maternal Hamilton Depression score. Points are colored by cell type (Hofbauer = light red, nRBC = dark yellow, Endothelial = dark red, Stromal = light yellow, Troph = dark blue, Syncytio = light blue).. SSRI = selective serotonin reuptake inhibitor, nRBC = nucleated red blood cells, Troph = non-multinucleated trophoblasts (primarily cytotrophoblasts), Syncytio = syncytiotrophoblast cell proportions.

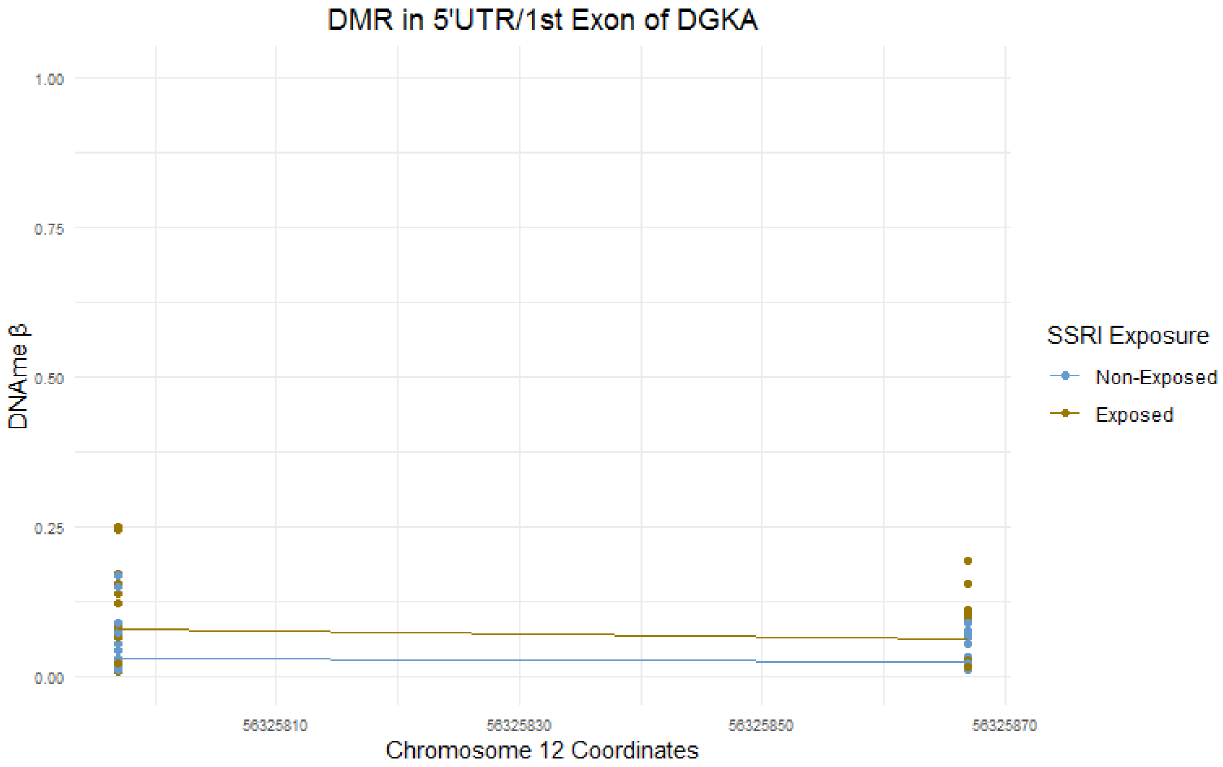


**Supplementary Figure 4. Differentially methylated region (DMR) associated with SSRI exposure in 5'UTR and 1st exon of *DGKA*.** Chromosome 12 hg19 coordinates (56,325,797-56,325,867) plotted along the X axis, DNAme β values at both CpGs (cg14921691, cg06762403) are plotted along the Y axis; points are colored by colored by SSRI exposure (dark yellow = SSRI-exposed, blue = non-SSRI-exposed). Average Δβ across this DMR is +0.06 (higher in SSRI-exposed placentas), and the minimum smoothed FDR calculated by DMRcate across this region was $1.95\times10^{-29}$.

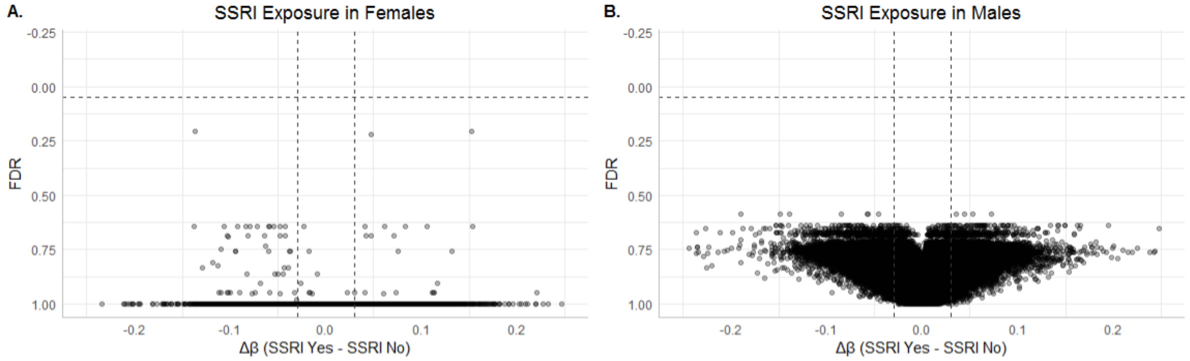


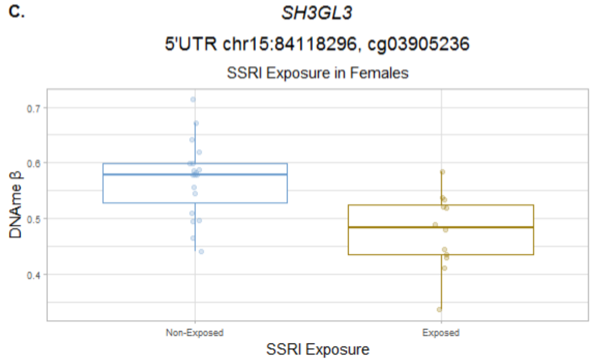


D.

| CpG | FDR | Δβ | Pos | Gene | Relation to Gene |
|---|---|---|---|---|---|
| cg15849349 | 0.21 | 0.15 | chr4:60419633 | -- | |
| cg07481545 | 0.22 | 0.05 | chr6:34131438 | -- | |
| cg03905236 | 0.21 | -0.13 | chr15:84118296 | *SH3GL3* | 5'UTR |

**Supplementary Figure 5. SSRI exposure is not associated with widespread DNAme patterns in sex-stratified subsets of the discovery cohort. (A)** Volcano plot showing the association between DNAme in female placentae and SSRI exposure, and **(B)** Volcano plot showing the association between DNAme in male placentae and SSRI exposure. For both plots SSRI refers to selective serotonin reuptake inhibitors, the difference in DNAme (Δβ) is plotted along the X axis and was calculated as $\Delta\beta = \beta_{\text{SSRI-exposed}} - \beta_{\text{SSRI non-exposed}}$, FDR is shown along the Y axis with more significant (lower FDR) values at the top of the plot. Vertical dashed intercepts demarcate $\Delta\beta = \pm 0.03$, a horizontal dashed intercept indicates FDR = 0.05. **(C)** Boxplot of differential DNAme by SSRI exposure status in female samples only (n=31) at cg03905236, in the 5' untranslated region (UTR) of the *SH3GL3* gene on chromosome 15. Points are colored by SSRI exposure (blue = SSRI non-exposed, dark yellow = SSRI-exposed) and boxplots indicate mean DNAme β value ± one standard deviation. **(D)** Table of CpGs with differential DNAme at FDR < 0.25 by SSRI exposure status in females only (n=31). Δβ refers to the difference in DNAme between groups and was calculated as $\Delta\beta = \beta_{\text{SSRI-exposed}} - \beta_{\text{SSRI non-exposed}}$. Pos refers to the genomic coordinates of the CpG in hg19.

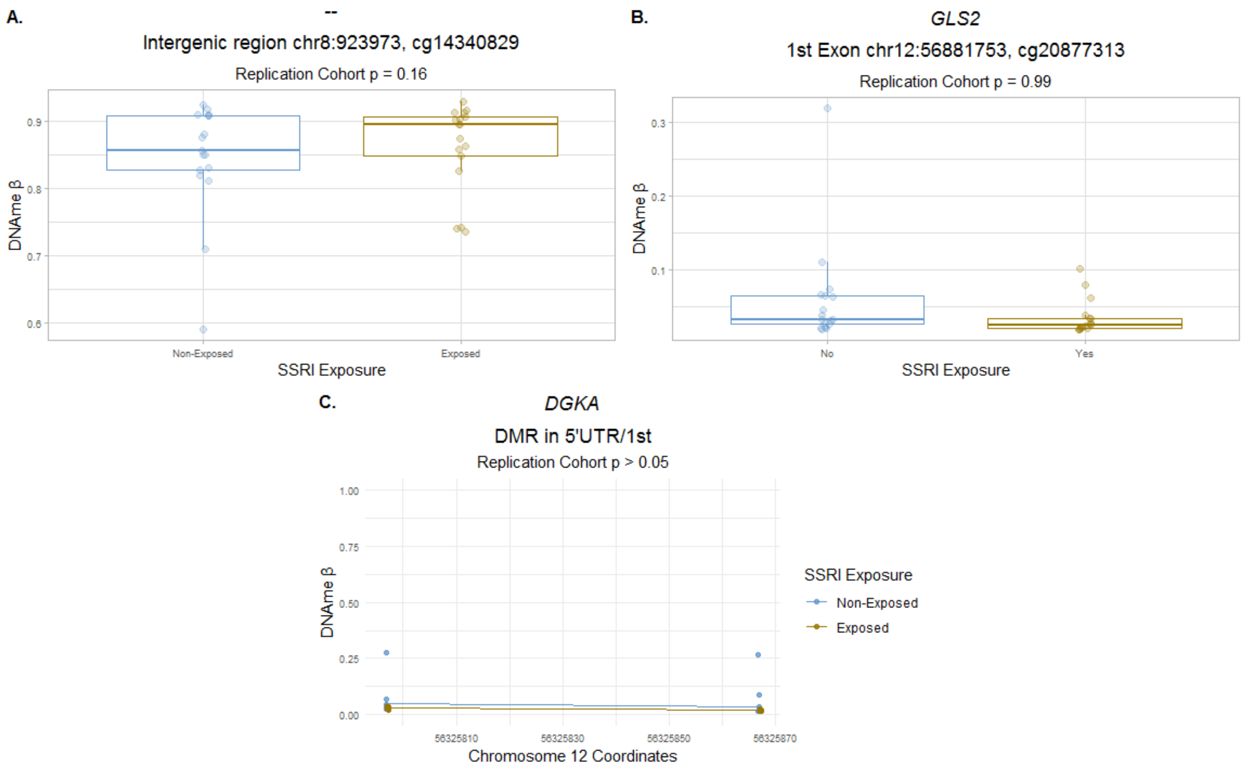


**Supplementary Figure 6. Testing discovery cohort differentially methylated CpGs for replication in GSE75248. (A)** Boxplot of mean DNAme β values ± one standard deviation in SSRI-exposed versus SSRI non-exposed samples at cg14340829 in the replication cohort; points are colored by SSRI exposure (blue = SSRI non-exposed, dark yellow = SSRI-exposed). **(B)** Boxplot of mean DNAme β values ± one standard deviation in SSRI-exposed versus SSRI non-exposed samples at cg20877313 in the replication cohort; points are colored by SSRI exposure (blue = SSRI non-exposed, dark yellow = SSRI-exposed). **(C)** Plot of DNAme β values in the replication cohort by SSRI exposure status in the SSRI-associated differentially methylated region in the *DGKA* gene, points are colored by SSRI exposure (blue = SSRI non-exposed, dark yellow = SSRI-exposed)

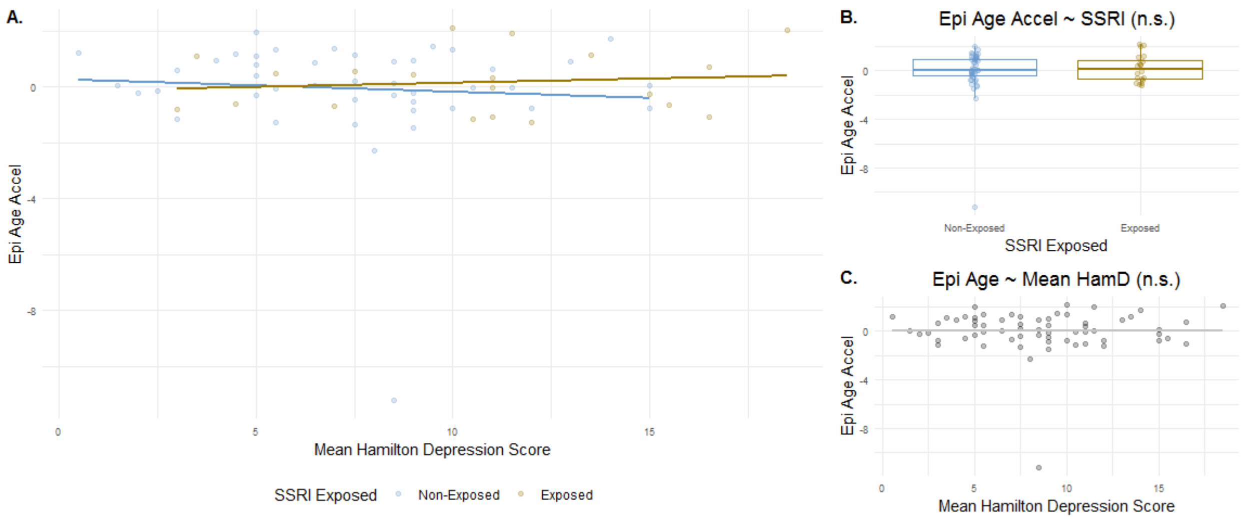


**Supplementary Figure 7. Placental epigenetic age acceleration is not associated with SSRI exposure or mean maternal Hamilton Depression score. (A)** Placental epigenetic age acceleration (Epi Age Accel) versus mean Hamilton Depression (HamD) score across gestation, colored by SSRI exposure (blue = SSRI non-exposed, dark yellow = SSRI-exposed). **(B)** Boxplot of placental epigenetic age acceleration separated by SSRI exposure status; boxplots indicate mean ± one standard deviation. **(C)** Scatterplot of mean Hamilton depression score versus placental epigenetic age acceleration with a line of best fit.

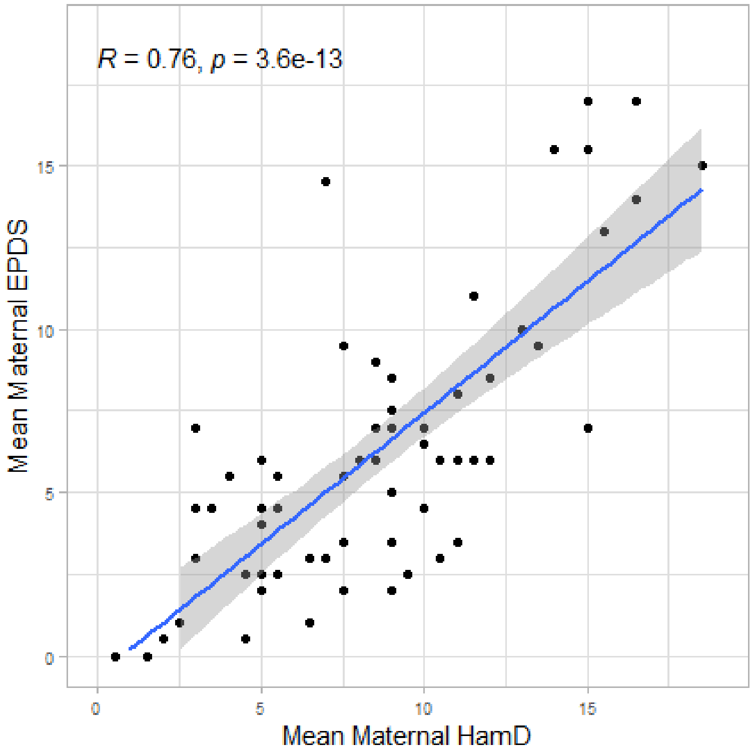


**Supplementary Figure 8. Mean maternal Hamilton Depression Score is significantly correlated with mean maternal Edinburgh Postnatal Depression Score in the discovery cohort.** Mean maternal Hamilton Depression score (HamD) is plotted along the X axis, mean

Edinburgh Postnatal Depression score (EPDS) is plotted along the Y axis, these depression metrics have a significant Pearson correlation coefficient of 0.76.

#### Supplementary Tables

**Supplementary Table 1. Results (number of significant CpGs) from SSRI exposure and maternal depression differential DNAme analyses.** Model A refers to linear modelling to identify DNAme alterations associated with SSRI exposure in full cohort (n=64), adjusting for mean maternal Hamilton Depression score across gestation. Model B refers to linear modelling to identify DNAme alterations associated with SSRI exposure in depressed samples (n=34, all mean maternal Hamilton Depression score > 8). Model C refers to linear modelling to identify DNAme alterations associated with maternal depression in SSRI non-exposed samples (n=44).

| Model | Threshold | $\|\Delta\beta\| > 0.00$ | $\|\Delta\beta\| > 0.03$ |
|---|---|---|---|
| A | FDR < 0.05 | 0 | 0 |
| | FDR < 0.15 | 1 | 1 |
| | FDR < 0.25 | 2 | 2 |
| B | FDR < 0.05 | 0 | 0 |
| | FDR < 0.15 | 3 | 3 |
| | FDR < 0.25 | 3 | 3 |
| C | FDR < 0.05 | 0 | 0 |
| | FDR < 0.15 | 0 | 0 |
| | FDR < 0.25 | 0 | 0 |
